# The association of umbilical cord blood neurofilament light with non-reassuring fetal status: a prospective observational study

**DOI:** 10.1101/2025.01.23.25320706

**Authors:** David Zalcberg, Kaitlin Kramer, Emma Payne, Thomas Payne, Shreeya Marathe, Neha Mahajan, Ashly Liu, Jessica Barry, Andrew Duckworth, Mitchell Brookes, Bradley de Vries, Fernando Gonzalez-Ortiz, Kaj Blennow, Henrik Zetterberg, Adrienne Gordon, Benjamin Moran, Helen Manning, Robert D. Sanders

## Abstract

**Objective:** Early detection of hypoxic–ischaemic encephalopathy (HIE) in neonates is critical. We conducted a pilot cohort study to determine the feasibility of collecting umbilical cord blood samples for neurofilament light (NfL) and to assess the association of NfL with non-reassuring fetal status and other cord biomarkers.

**Design:** Prospective cohort study.

**Setting:** A single, large tertiary and quaternary referral hospital.

**Patients:** 108 maternal participants consenting to donate cord blood.

**Intervention:** Umbilical cord venous blood plasma NfL levels.

**Main outcome measures:** (1) Feasibility of cord NfL sample collection and analysis; (2) Association of NfL with non-reassuring fetal status (CTG changes and/or documented non-reassuring fetal status), NICU admission and length of stay; (3) Correlation of NfL with other cord biomarkers.

**Results:** Cord NfL was higher in preterm neonates, and was correlated with cord lactate, pH, and base excess. After controlling for mode of delivery and gestational age, NfL (OR = 2.29, 95%CI: 1.15 to 5.57), but not pH (OR = 0.78, 95%CI: 0.42 to 1.41), base excess (OR = 0.83, 95%CI: 0.37 to 1.86), or lactate (OR = 1.06, 95%CI: 0.51 to 2.12) was associated with non-reassuring fetal status. NfL levels were higher in neonates admitted to NICU (median (IQR): 11.3 (7) versus 8.5 (5.1)).

**Conclusions:** Cord blood NfL analysis was feasible and provided correlates of adverse outcomes. Higher venous cord blood NfL levels were associated with non-reassuring fetal status. Further research is needed to validate these findings and establish the role of NfL, if any, in clinical practice.

**What is already known on this topic:** Umbilical cord blood NfL is a promising biomarker of neuronal injury in neonates with overt HIE. Whether cord NfL may improve diagnosis of mild–moderate HIE via identification of in utero hypoxia is unknown.

**What this study adds:** We found that cord NfL is associated with non-reassuring fetal status, and outperforms other cord biomarkers.

**How this study might affect research, practice, or policy:** This study lays the groundwork for future research into the use of cord NfL for HIE diagnosis and risk stratification. It supports ongoing development of a point-of-care scalp NfL assay.

## Introduction

Hypoxic–ischaemic encephalopathy (HIE) is a common cause of neonatal encephalopathy (NE), occurring in 1–2 per 1000 live births.^1^ Therapeutic hypothermia is the only proven treatment for HIE,^2–5^ and must be started within 6 hours of birth.^1^ Current practice for identifying HIE relies on clinical signs, including need for resuscitation and cord blood acid– base analysis, which has limited sensitivity in part because cord blood acid–base biomarkers are non-specific for detection of neuronal injury.^6, 7^ Identification of in utero fetal hypoxia can help to direct expedited delivery of the fetus, in an attempt to decrease the incidence of HIE. A composite outcome ‘non-reassuring fetal status’ may be used as an indicator of fetal hypoxia, and is defined by abnormalities in fetal heart rate (primarily detected by cardiotocography (CTG)), fetal scalp blood sampling, and ultrasound parameters.^8, 9^

Neurofilament light (NfL) is a neuronal protein released at the time of many types of neuronal injuries.^10^ Two case–control studies of cord NfL have suggested associations with HIE and perinatal morbidity.^11, 12^ However, research is lacking regarding the correlation between cord NfL and non-reassuring fetal status.^13^ Demonstration of such an association would reinforce the use of NfL as a possible adjunct in the diagnosis of HIE, and support continued development of a rapid point-of-care scalp NfL test that may, in future, help identify in utero hypoxia.

We conducted a prospective cohort study to determine the association of umbilical cord NfL with various perinatal factors. Our hypotheses were:

1. Collection and analysis of cord blood NfL is feasible;
2. Cord NfL has a stronger association with non-reassuring fetal status than other cord biomarkers; and
3. Cord NfL is correlated with other cord markers of non-reassuring fetal status.

## Methods

### Study Design

A prospective cohort study was conducted with women who birthed at Royal Prince Alfred Hospital (RPAH). RPAH is an inner-city tertiary and quaternary public referral hospital in Sydney with mostly urban-dwelling patients. Participants were prospectively recruited at any time from 4 weeks prior to estimated delivery date, to date of delivery (including during delivery). Informed consent was obtained from the birth parent for all participants. Study data were collected and stored in REDCap by the research team. Ethics approval was obtained from Sydney Local Health District Human Research and Ethics Committee (approval number: 2022/ETH01100). Data were reported in accordance with the Strengthening the Reporting of Observational Studies in Epidemiology (STROBE) guidelines.^14^

### Study Population

Women who birthed at RPAH from October 2022 were eligible for study recruitment, including all live births delivered vaginally or via caesarean section (CS). Women were excluded if they were non-English speaking or had a history of psychological illness, or other conditions, that interfered with capacity to provide informed consent.

### Biospecimen Collection and Analysis

Immediately at birth, 3mL of umbilical venous cord blood was collected in EDTA tubes from each study participant. Blood samples were centrifuged in the Department of Anaesthetics laboratory, and plasma aliquots stored in barcoded cryovials.

Plasma samples were sent to the University of Gothenburg in Sweden for analysis. Plasma NfL concentration was measured using the NF-Light assay on a Single molecule array (Simoa) HD-X instrument according to instructions from the manufacturer (Quanterix, Billerica, MA). All measurements were done with a four-fold dilution factor in singlicates and performed on one occasion using one batch of reagents with the analyst blinded to clinical data. Intra-assay coefficients of variation were <10% derived from internal control plasma samples measured in duplicate on each analytical run.

### Demographic and Outcome Data Collection

Demographic and outcome data were extracted from electronic medical records. Data extraction included antenatal history, maternal and child health factors, intrapartum events and monitoring, adverse perinatal events, and any health events up to four weeks postpartum. Researchers collecting these data were blinded to the NfL results.

### Outcomes

#### Primary trial feasibility outcomes

Our primary feasibility outcomes were the proportion of eligible participants recruited to the study and the proportion of enrolled participants who had cord blood samples successfully sent for analysis.

#### Primary clinical outcome

Our primary clinical outcome was the association of cord NfL with non-reassuring fetal status. ‘Non-reassuring fetal status’ was defined as any record of non-reassuring fetal status in the intrapartum or antenatal record of care, as well as any documented cardiotocography (CTG) changes aligning with ‘red zone’ criteria per the NSW Health electronic fetal monitoring guidelines (Appendix 1). We adjusted our primary outcome for mode of delivery (vaginal or elective CS or emergency CS) and gestational age, given previous work has suggested that vaginal delivery and longer gestation are associated with greater cord NfL, and we hypothesised that these would also be associated with non-reassuring fetal status.^13^

#### Secondary clinical outcomes

Secondary outcomes included the association of NfL with other cord biomarkers (lactate, base excess, pH), intrapartum factors (mode of delivery, duration of labour), fetal factors (gestational age, head circumference, birthweight), and other neonatal outcomes (resuscitation requirement, NICU admission, NICU length of stay). A full list is provided in Appendix 2. We *a priori* defined preterm as birth at <37 weeks’ gestation and very preterm as <32 weeks’ gestation. We defined resuscitation requirement as any continuous positive airway pressure requirement postnatally. Analyses of birthweight and head circumference were adjusted for gestational age, as this has been suggested to be a confounder.^13^ While no previous work exists, we hypothesised that analyses of resuscitation requirement required adjustment for gestational age, while analyses of NICU length of stay required adjustment for mode of delivery and gestational age.

### Power analysis

Previous work has suggested the standard deviation (SD) of umbilical cord NfL is 13 pg/mL.^11^ To ensure that the 95% confidence interval estimate for mean NfL in our sample was within 3 pg/mL of the true population mean, 73 neonates were required. Anticipating possible issues with sample storage/shipment, 110 mothers were recruited. Using the previously reported incidence of non-reassuring fetal status of approximately 16%,^8^ 110 participants yields 80% power to detect a large effect size (Cohen’s d = 0.8), based on a two-sided *t*-test with α = 0.025 (halved from 0.05 to account for our dual feasibility/clinical primary outcome).

### Statistical methods

NfL concentrations showed a strong positive skew, so were log_10_ transformed for all analyses. Where the dependent variable was binary, we used logistic regression. Biomarker concentrations were standardised in these models by subtracting the mean and dividing by the standard deviation. Area under the receiver operator curve (AUROC) was calculated for biomarkers in predicting non-reassuring fetal status. The 95% confidence interval for the AUROC was calculated using 2000 stratified bootstrap replicates. The calculation of Youden’s Index from the receiver–operator curve (ROC)^15^ was used to determine a binary cutoff value for all biomarkers that maximised the sum of the specificity and sensitivity. We did not undertake model calibration nor perform cross-validation—our analyses aimed to be descriptive.

Regarding secondary outcomes, we report rank-based nonparametric methods for bivariate biomarker analysis. Linear regression with ordinary least squares estimation was used for multivariable models where pH or base excess was the dependent variable. Where log_10_ NfL or lactate was the dependent variable, we used a generalised linear model with an inverse Gaussian family with log link. We aimed to model count data using the simplest possible distribution (the Poisson distribution), unless overdispersion was present, in which case we used a negative binomial family model. Nested models were compared using F-tests (or χ^2^ tests for count models). As this study was hypothesis-generating, no adjustments were made for multiple comparisons.

All analyses were conducted in R using RStudio (Version 2023.06.1; R Foundation for Statistical Computing, Vienna, Austria), using the ‘lme4’, ‘plotROC’, and ‘pROC’ packages.

## Results

Cohort demographic information is summarised in Supplementary Table 1.

### Primary outcomes

#### Feasibility of testing cord NfL

Of 509 births at RPAH over the period 24th October 2022 to 9th of December 2022, 110 women were recruited, meaning 22% of all eligible participants were enrolled in the study. A total of 108 participants had cord blood sent for analysis (STROBE diagram: Supplementary Figure 1), representing a 98% conversion rate from enrolment to analysis.

#### Association of neurofilament light with non-reassuring fetal status

Participants classified as experiencing non-reassuring fetal status (n = 16) had higher cord NfL (median (Q1, Q3) NfL: 14.8 (11.2, 18.0) vs. 8.4 (6.7, 11.4)). These participants also experienced higher lactate levels, as well as lower cord pH and base excess (Supplementary Figure 2). The association between log_10_ NfL (in standardised form) and non-reassuring fetal status persisted after adjusting for mode of delivery and gestational age (adjusted OR per 1 SD increase in log_10_ NfL = 2.29, 95%CI: 1.15 to 5.57; p = 0.038), whereas pH, base excess, and lactate were not associated with non-reassuring fetal status in the adjusted model (Table 1).

**Table 1.** Results from binomial-family models predicting non-reassuring fetal status. Results for all four biomarkers are shown. Biomarkers were standardised by subtracting the mean and dividing by the standard deviation. AIC = Akaike information criterion; BIC = Bayesian information criterion; NfL = neurofilament light.

| Characteristic | Log10 cord NfL<sup>1</sup> |  |  | Cord pH<sup>2</sup> |  |  | Cord base excess<sup>3</sup> |  |  | Cord lactate<sup>4</sup> |  |  |
| --- | --- | --- | --- | --- | --- | --- | --- | --- | --- | --- | --- | --- |
|  | OR<sup>5</sup> | 95% CI<sup>5</sup> | p-value<sup>6</sup> | OR<sup>5</sup> | 95% CI<sup>5</sup> | p-value<sup>6</sup> | OR<sup>5</sup> | 95% CI<sup>5</sup> | p-value<sup>6</sup> | OR<sup>5</sup> | 95% CI<sup>5</sup> | p-value<sup>6</sup> |
| **Unadjusted model** |  |  |  |  |  |  |  |  |  |  |  |  |
| Cord biomarker (standardised) | 2.20 | 1.35, 4.00 | 0.004** | 0.49 | 0.27, 0.85 | 0.013* | 0.40 | 0.21, 0.70 | 0.002** | 2.19 | 1.33, 3.74 | 0.003** |
| **Model adjusted for mode of delivery and gestational age** |  |  |  |  |  |  |  |  |  |  |  |  |
| Cord biomarker (standardised) | 2.29 | 1.15, 5.57 | 0.038* | 0.78 | 0.42, 1.41 | 0.423 | 0.83 | 0.37, 1.86 | 0.650 | 1.06 | 0.51, 2.12 | 0.862 |
| <sup>1</sup> Unadjusted model: AIC = 79.8; BIC = 85.1; No. Obs. = 105. Adjusted model: AIC = 70.1; BIC = 83.3; No. Obs. = 105; p-value from F-test comparing unadjusted vs. adjusted model fit = 0.001<br><sup>2</sup> Unadjusted model: AIC = 79.7; BIC = 85.0; No. Obs. = 104. Adjusted model: AIC = 70.2; BIC = 83.5; No. Obs. = 104; p-value from F-test comparing unadjusted vs. adjusted model fit = 0.001<br><sup>3</sup> Unadjusted model: AIC = 65.7; BIC = 70.8; No. Obs. = 94. Adjusted model: AIC = 62.1; BIC = 74.8; No. Obs. = 94; p-value from F-test comparing unadjusted vs. adjusted model fit = 0.023<br><sup>4</sup> Unadjusted model: AIC = 76.6; BIC = 81.9; No. Obs. = 103. Adjusted model: AIC = 70.8; BIC = 84.0; No. Obs. = 103; p-value from F-test comparing unadjusted vs. adjusted model fit = 0.008<br><sup>5</sup> OR = Odds Ratio, CI = Confidence Interval<br><sup>6</sup> *p<0.05; **p<0.01; ***p<0.001 |  |  |  |  |  |  |  |  |  |  |  |  |

The combination of NfL with delivery mode (vaginal or elective CS or emergency CS) and gestational age provided a slightly better AUC (0.87 95%CI: 0.79 to 0.95) for the association with non-reassuring fetal status than the combination of gestational age and delivery mode with the other biomarkers (Figure 1A). When using biomarkers alone for prediction of non-reassuring fetal status, the most pronounced difference between NfL and other biomarkers was seen when excluding elective CS and preterm births (Figure 1F).

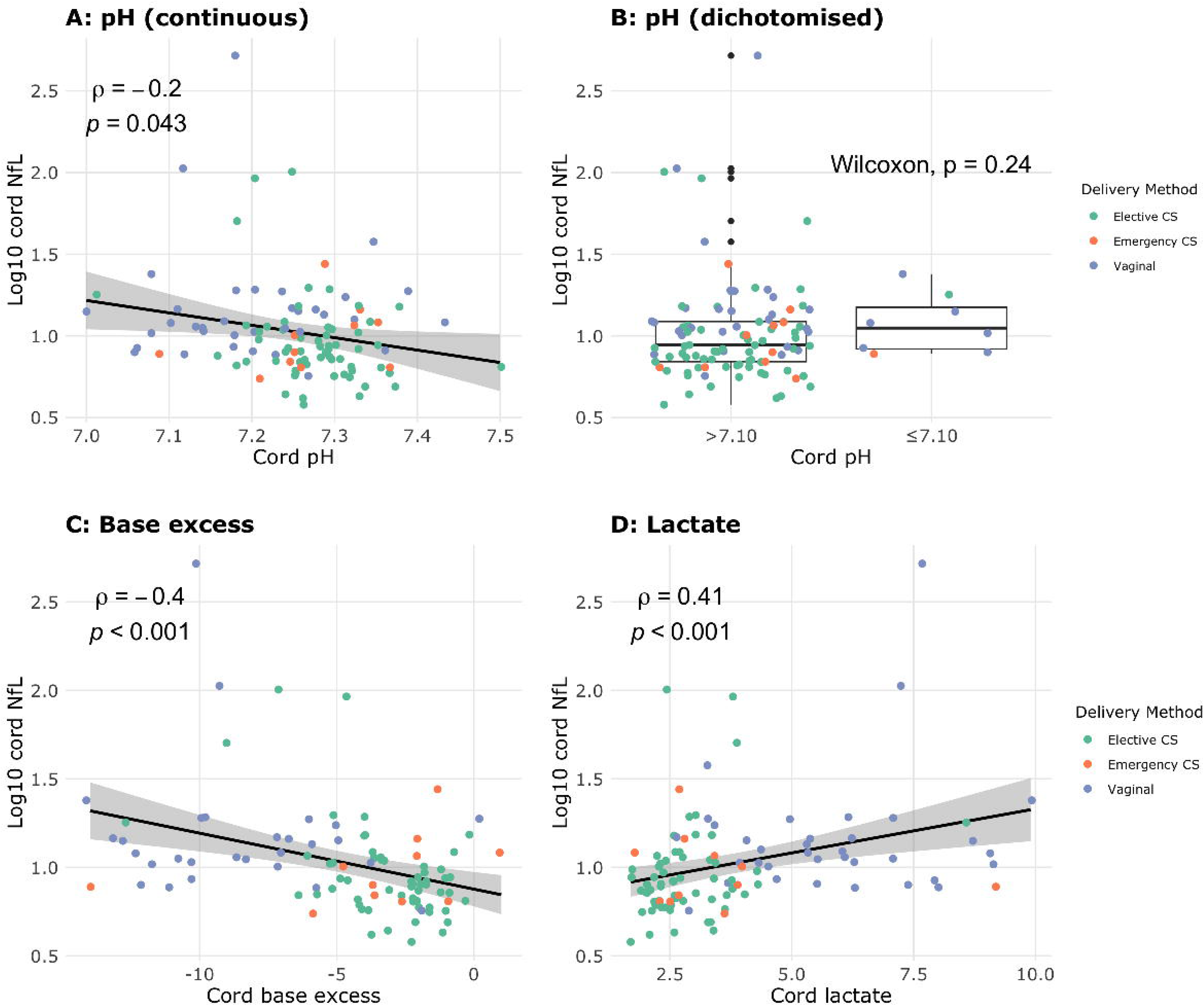

Youden’s Index was calculated for all biomarkers. When including all births, diagnostic test characteristics of log_10_ NfL >1.09 included a negative predictive value of 0.96 (Table 2). In general, the specificity and positive predictive value of various log_10_ NfL cutoffs was much better than other biomarkers, while sensitivity was lower.

**Table 2.** Test characteristics of cord biomarkers in predicting non-reassuring fetal status. The cutoff value is determined by the calculation of Youden’s Index from the receiver–operator curve, which maximises the sum of the sensitivity + specificity. CS = caesarean section; NfL = neurofilament light.

|  | **Threshold**<br>**(95%CI)<sup>1</sup>** | **Sensitivity**<br>**(95%CI)** | **Specificity**<br>**(95%CI)** | **Positive**<br>**predictive**<br>**value**<br>**(95%CI)** | **Negative**<br>**predictive**<br>**value**<br>**(95%CI)** |
| --- | --- | --- | --- | --- | --- |
| **All births** |  |  |  |  |  |
| Log10 NfL | 1.09 (0.90, 1.17) | 0.80 (0.60, 1.00) | 0.84 (0.43, 0.92) | 0.44 (0.23, 0.64) | 0.96 (0.92, 1.00) |
| pH | 7.25 (7.13, 7.31) | 0.79 (0.43, 1.00) | 0.66 (0.26, 0.91) | 0.27 (0.17, 0.50) | 0.95 (0.91, 1.00) |
| Base excess | -4.50 (-6.50, -3.50) | 1.00 (0.75, 1.00) | 0.63 (0.45, 0.83) | 0.28 (0.21, 0.42) | 1.00 (0.96, 1.00) |
| Lactate | 3.95 (3.35, 5.00) | 0.86 (0.64, 1.00) | 0.75 (0.51, 0.88) | 0.34 (0.23, 0.50) | 0.97 (0.93, 1.00) |
| **Excluding preterm births** |  |  |  |  |  |
| Log10 NfL | 1.09 (0.90, 1.16) | 0.77 (0.54, 1.00) | 0.86 (0.44, 0.95) | 0.48 (0.22, 0.72) | 0.96 (0.93, 1.00) |
| pH | 7.25 (7.17, 7.25) | 0.77 (0.46, 1.00) | 0.68 (0.45, 0.90) | 0.29 (0.20, 0.50) | 0.96 (0.91, 1.00) |
| Base excess | -4.50 (-6.50, -4.50) | 1.00 (0.82, 1.00) | 0.66 (0.52, 0.82) | 0.30 (0.24, 0.44) | 1.00 (0.97, 1.00) |
| Lactate | 3.95 (3.35, 5.00) | 0.85 (0.69, 1.00) | 0.76 (0.52, 0.87) | 0.37 (0.24, 0.52) | 0.97 (0.93, 1.00) |
| **Excluding elective CS births** |  |  |  |  |  |
| Log10 NfL | 1.12 (0.90, 1.17) | 0.71 (0.43, 1.00) | 0.79 (0.27, 0.97) | 0.58 (0.37, 0.87) | 0.86 (0.77, 1.00) |
| pH | 7.25 (7.12, 7.32) | 0.92 (0.46, 1.00) | 0.48 (0.18, 0.88) | 0.40 (0.33, 0.63) | 0.94 (0.79, 1.00) |
| Base excess | -4.50 (-11.50, -4.00) | 1.00 (0.45, 1.00) | 0.43 (0.25, 0.93) | 0.41 (0.34, 0.75) | 1.00 (0.81, 1.00) |
| Lactate | 4.45 (3.35, 7.25) | 0.85 (0.31, 1.00) | 0.52 (0.21, 0.94) | 0.41 (0.32, 0.71) | 0.89 (0.76, 1.00) |
| **Excluding elective CS and preterm births** |  |  |  |  |  |
| Log10 NfL | 1.11 (0.90, 1.22) | 0.69 (0.38, 1.00) | 0.82 (0.32, 1.00) | 0.65 (0.39, 1.00) | 0.85 (0.76, 1.00) |
| pH | 7.25 (7.08, 7.31) | 0.85 (0.31, 1.00) | 0.46 (0.11, 0.93) | 0.42 (0.34, 0.69) | 0.88 (0.73, 1.00) |
| Base excess | -9.50 (-Inf, Inf) | 0.73 (0.00, 1.00) | 0.46 (0.00, 1.00) | 0.36 (0.31, 0.50) | 0.76 (0.69, 1.00) |
| Lactate | 4.45 (2.94, 9.55) | 0.85 (0.08, 1.00) | 0.50 (0.11, 1.00) | 0.43 (0.33, 1.00) | 0.83 (0.70, 1.00) |

<sup>1</sup> Obtained using the calculation of Youden's Index (maximum sum of sensitivity + specificity).

### Secondary outcomes

Cord NfL demonstrated an inverse correlation with cord pH and base excess, and a positive correlation with cord blood lactate (Figure 2). Conversely, examining only participants who laboured, no association was found for any biomarker (Supplementary Figure 3).

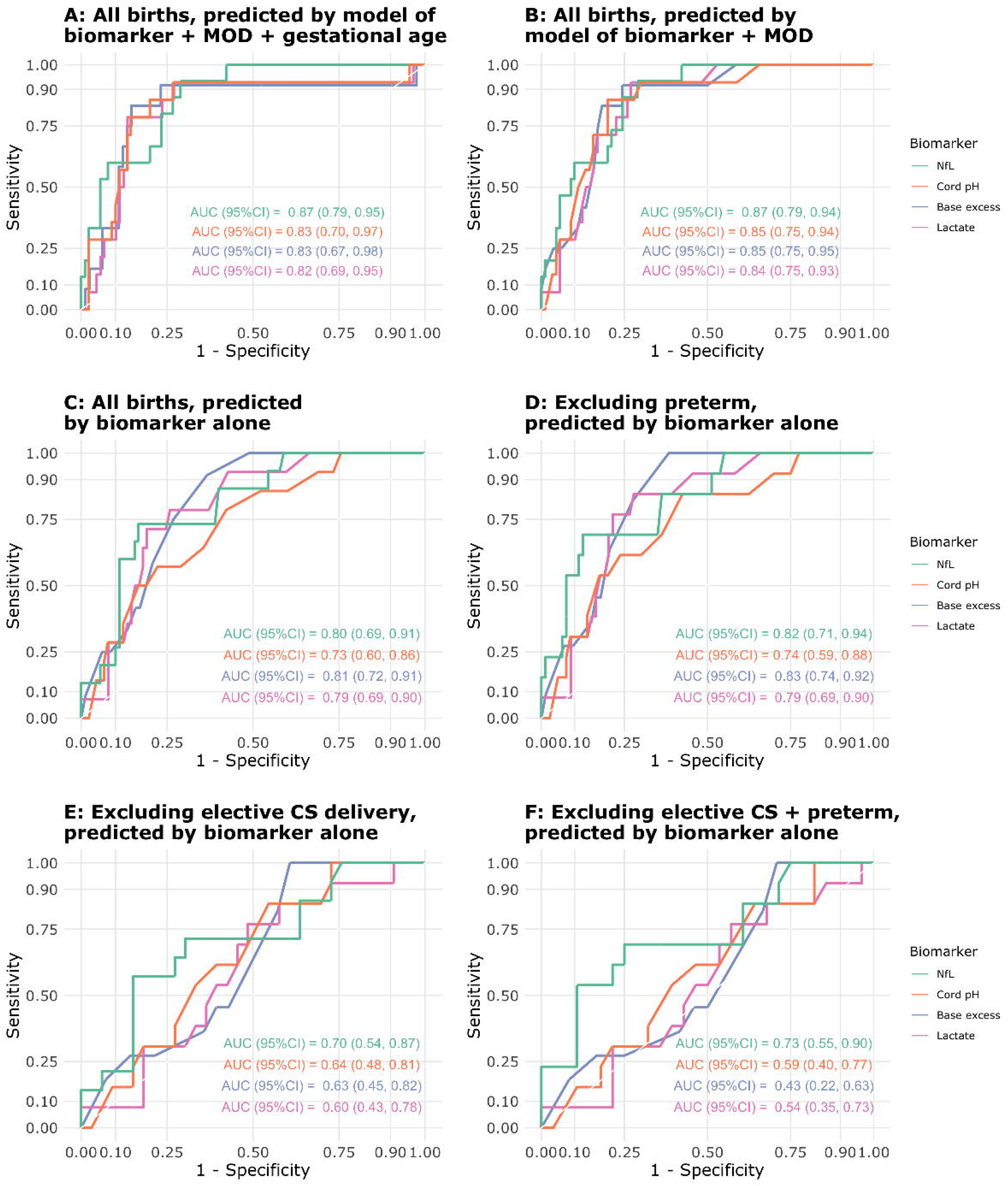

NfL, pH, base excess, and lactate values differed significantly by mode of delivery (Kruskal– Wallis, all p < 0.001) (Figure 3). There was no statistically significant correlation of any biomarker with duration of 1^st^ stage of labour. Whilst duration of the 2^nd^ stage was negatively correlated with pH (Spearman ρ = –0.49, p = 0.009) and base excess (Spearman ρ = –0.44, p = 0.044) and positively correlated with lactate (Spearman ρ = 0.40, p = 0.037), there was no statistically significant correlation with NfL (Spearman ρ = –0.27, p = 0.17) (Supplementary Figure 4).

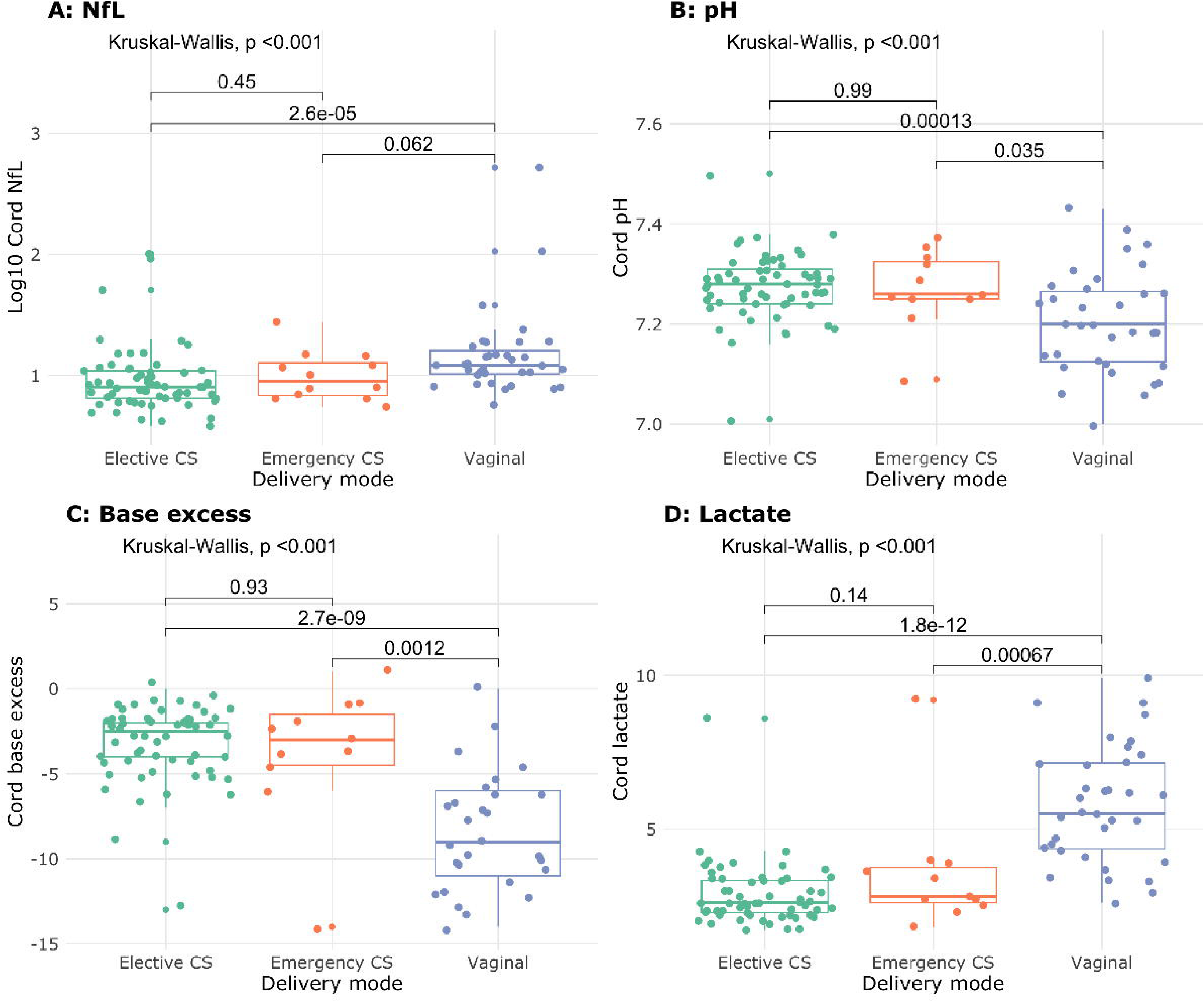

We observed a lower cord NfL level in participants who were born term (n = 94) compared to those born preterm (under 37 completed weeks gestation; n = 10), and very preterm (under 32 completed weeks gestation; n = 4) (median (Q1, Q3) log_10_ NfL = 0.94 (0.84, 1.08) vs. 1.18 (0.97, 1.48) vs. 1.17 (1.14, 1.24); Kruskal–Wallis p = 0.004) (Supplementary Figure 5). Conversely, we did not observe a difference in cord pH, base excess, or lactate across these groups of gestational age.

Keeping gestational age constant, every 1 kg increase in birth weight was associated with an 11% decrease in mean log_10_ NfL (exp(β) = 0.89, 95%CI: 0.81 to 0.98; p = 0.025) (Supplementary Table 2). There was no significant relationship with biomarkers and head circumference in adjusted analysis (Supplementary Table 3).

We did not observe evidence of an association of NfL, pH, base excess, or lactate with resuscitation requirement after adjusting for preterm birth (Supplementary Table 4). NfL values were significantly higher in participants who required NICU admission (median (Q1, Q3) log_10_ NfL: 1.05 (0.91, 1.18) vs. 0.93 (0.84, 1.08); Wilcoxon p = 0.040). Conversely, we did not observe evidence for a difference in cord pH and lactate values in neonates admitted to NICU vs. those not, while base excess values were slightly higher in those admitted to NICU (Supplementary Figure 6).

We did not observe strong evidence to suggest higher NfL values (in standardised form) were associated with an increased NICU length of stay in the unadjusted model (incidence rate ratio (IRR) = 3.04, 95%CI: 0.94 to 9.80; p = 0.063), or after adjusting for gestational age and mode of delivery (IRR = 1.51, 95%CI: 0.71 to 3.19; p = 0.284) (Supplementary Table 5). Cord pH, base excess, and lactate were all associated with NICU length of stay in the unadjusted models, but none were statistically significant in the adjusted model (Supplementary Table 5).

## Discussion

We observed an association between higher cord blood NfL with non-reassuring fetal status, which remained robust after adjusting for mode of delivery and gestational age. In addition, we showed that NfL correlates with cord biomarkers of neonatal asphyxia. Together, our findings provide valuable evidence of the associations between NfL and surrogates of in-utero hypoxia, which is supportive of future studies of NfL in the diagnosis and treatment of HIE.

The largest descriptive study of cord blood NfL to date (n = 665), by Kurner and colleagues, supports our findings of higher NfL levels in vaginal delivery.^13^ Meanwhile, in that study, higher gestational age was associated with greater cord NfL, while in our cohort, there was a negative association. In addition, our study found a negative association between NfL and birthweight, whilst Kurner and colleagues found no evidence of this association. A possible reason for these discrepancies is that Kurner and colleagues included only healthy term neonates,^13^ whilst we included a higher risk cohort (24% admitted to NICU), and 13% of our cohort were born preterm. NfL appears to be higher in preterm neonates, who also have lower birthweight.

Our study adds to previous research regarding the association between NfL and adverse outcomes in neonates. Studies of cord NfL in HIE are limited to case–control designs given the rarity of overt HIE.^11, 12^ The two case–control studies (n = 38 and 150) of cord blood demonstrated a higher NfL in cases of asphyxia with or without HIE,^11, 12^ with asphyxia defined as Apgar ≤7 at 5⍰min and/or umbilical cord blood acidosis (pH <7.0). By linking cord NfL with non-reassuring fetal status, we attempt to bridge the results of these case–control studies to a possible role of NfL in the identification of in utero hypoxia, which may in future lead to studies of NfL in the diagnosis of milder or unrecognised cases of HIE. ‘Pathological’ fetal heart rate abnormalities, detected by CTG and ultrasound, may indicate early compensatory changes to hypoxia,^16^ and correlate with neonatal acidosis.^17–19^ Other studies of the plasma and CSF obtained in the days following birth have shown associations of NfL with neonatal asphyxia and HIE,^20, 21^ providing mechanistic support for our work.

Limitations of this study included a relatively small sample size (n = 108). In addition, our population included a high proportion of elective CS, which are excluded from intrapartum identification of non-reassuring fetal status by our criteria. In addition, we did not assess whether hypoxia suggested by non-reassuring fetal status led to downstream neurological effects. There is not yet evidence that increased intrapartum monitoring and identification of non-reassuring fetal status leads to decreased incidence of HIE overall, despite correlating with a reduction in neonatal seizure incidence.^22–25^ Moreover, fetal heart rate patterns have differing clinical implications depending on fetal and maternal background, and therefore having variable sensitivity and specificity for hypoxic injury.

## Conclusion

In conclusion, our study underscores the potential of NfL as a promising biomarker of non-reassuring fetal status, and hence fetal hypoxia.

## Details of author contributions

DZ and RDS designed the study in consultation with BdV, KK, BM, and HM. FG, HZ and KB supplied the assays and managed biofluid analysis. EP and TP conducted the statistical analysis. EP and DZ drafted the manuscript. All authors provided critical feedback on the manuscript.

## Funding

The work is supported by US National Institutes of Health (NIH) grant R01 AG063849-01 (RDS). KB is supported by the Swedish Research Council (#2017-00915 and #2022-00732), the Swedish Alzheimer Foundation (#AF-930351, #AF-939721, #AF-968270, and #AF-994551), Hjärnfonden, Sweden (#FO2017-0243 and #ALZ2022-0006), the Swedish state under the agreement between the Swedish government and the County Councils, the ALF-agreement (#ALFGBG-715986 and #ALFGBG-965240), the European Union Joint Program for Neurodegenerative Disorders (JPND2019-466-236), the Alzheimer’s Association 2021 Zenith Award (ZEN-21-848495), the Alzheimer’s Association 2022-2025 Grant (SG-23-1038904 QC), La Fondation Recherche Alzheimer (FRA), Paris, France, and the Kirsten and Freddy Johansen Foundation, Copenhagen, Denmark. HZ is a Wallenberg Scholar and a Distinguished Professor at the Swedish Research Council supported by grants from the Swedish Research Council (#2023-00356; #2022-01018 and #2019-02397), the European Union’s Horizon Europe research and innovation programme under grant agreement No 101053962, Swedish State Support for Clinical Research (#ALFGBG-71320), the Alzheimer Drug Discovery Foundation (ADDF), USA (#201809-2016862), the AD Strategic Fund and the Alzheimer’s Association (#ADSF-21-831376-C, #ADSF-21-831381-C, #ADSF-21-831377-C, and #ADSF-24-1284328-C), the Bluefield Project, Cure Alzheimer’s Fund, the Olav Thon Foundation, the Erling-Persson Family Foundation, Stiftelsen för Gamla Tjänarinnor, Hjärnfonden, Sweden (#FO2022-0270), the European Union’s Horizon 2020 research and innovation programme under the Marie Skłodowska-Curie grant agreement No 860197 (MIRIADE), the European Union Joint Programme – Neurodegenerative Disease Research (JPND2021-00694), the National Institute for Health and Care Research University College London Hospitals Biomedical Research Centre, and the UK Dementia Research Institute at UCL (UKDRI-1003).

## Supplementary material

Supplementary material is available online.

## Declarations of interests

KB has served as a consultant and at advisory boards for Acumen, ALZPath, AriBio, BioArctic, Biogen, Eisai, Lilly, Moleac Pte. Ltd, Novartis, Ono Pharma, Prothena, Roche Diagnostics, and Siemens Healthineers; has served at data monitoring committees for Julius Clinical and Novartis; has given lectures, produced educational materials and participated in educational programs for AC Immune, Biogen, Celdara Medical, Eisai and Roche Diagnostics; and is a co-founder of Brain Biomarker Solutions in Gothenburg AB (BBS), which is a part of the GU Ventures Incubator Program, outside the work presented in this paper.

HZ has served at scientific advisory boards and/or as a consultant for Abbvie, Acumen, Alector, Alzinova, ALZPath, Amylyx, Annexon, Apellis, Artery Therapeutics, AZTherapies, Cognito Therapeutics, CogRx, Denali, Eisai, Merry Life, Nervgen, Novo Nordisk, Optoceutics, Passage Bio, Pinteon Therapeutics, Prothena, Red Abbey Labs, reMYND, Roche, Samumed, Siemens Healthineers, Triplet Therapeutics, and Wave, has given lectures in symposia sponsored by Alzecure, Biogen, Cellectricon, Fujirebio, Lilly, and Roche, and is a co-founder of Brain Biomarker Solutions in Gothenburg AB (BBS), which is a part of the GU Ventures Incubator Program (outside submitted work). The other authors have no conflicts of interest to declare.

## Data availability

The raw data are available from the corresponding author upon reasonable request.

## Notes

### Author Declarations

Sydney Local Health District Human Research and Ethics Committee (approval number: 2022/ETH01100).

### Summary of Updates

Supplemental files updated for clarification

