## Supplementary for "The association of umbilical cord blood neurofilament light with non-reassuring fetal status: a prospective observational study"

**Supplementary Materials**

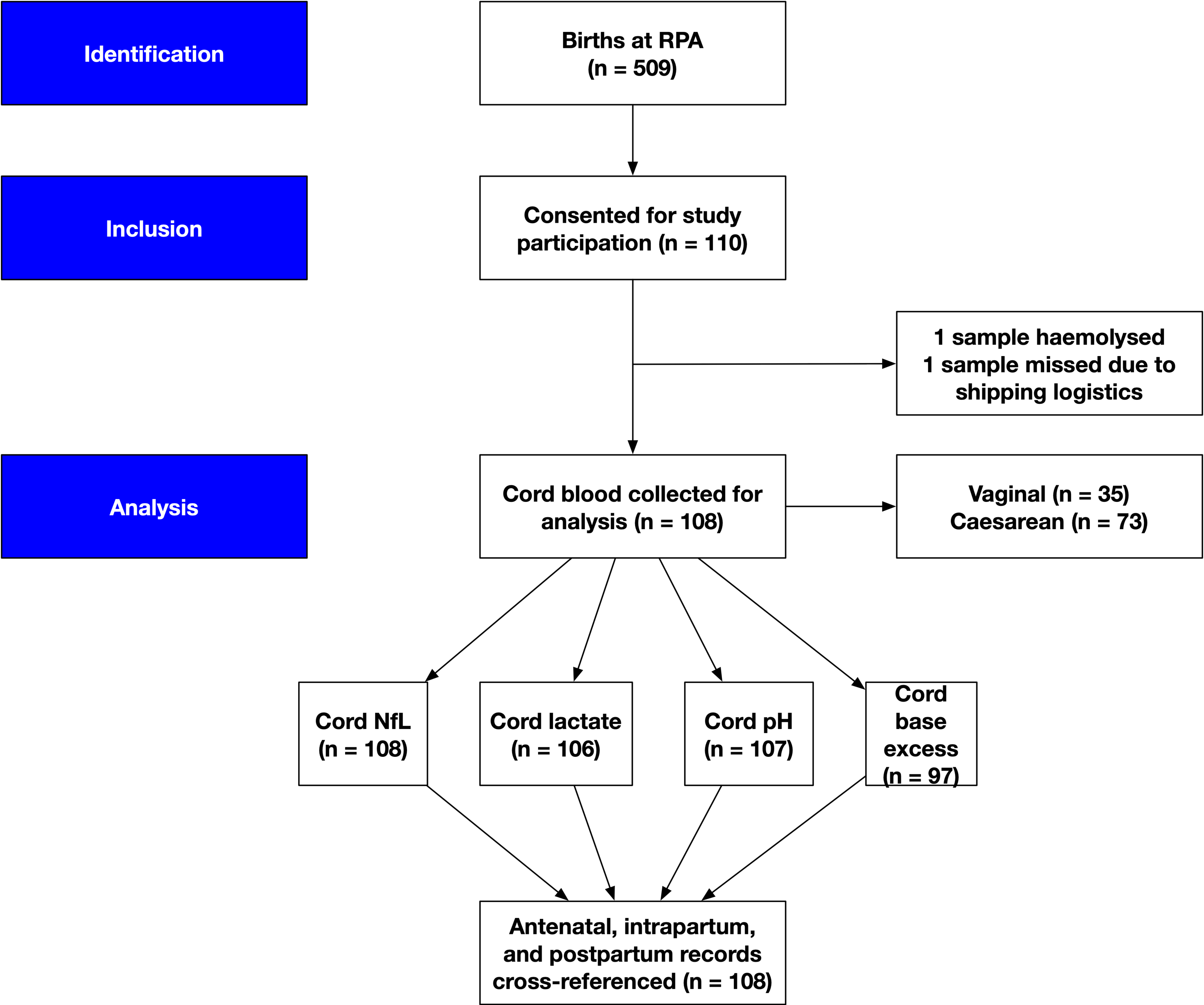
**Supplementary Figure 1**: STROBE diagram of participant inclusion for the study.

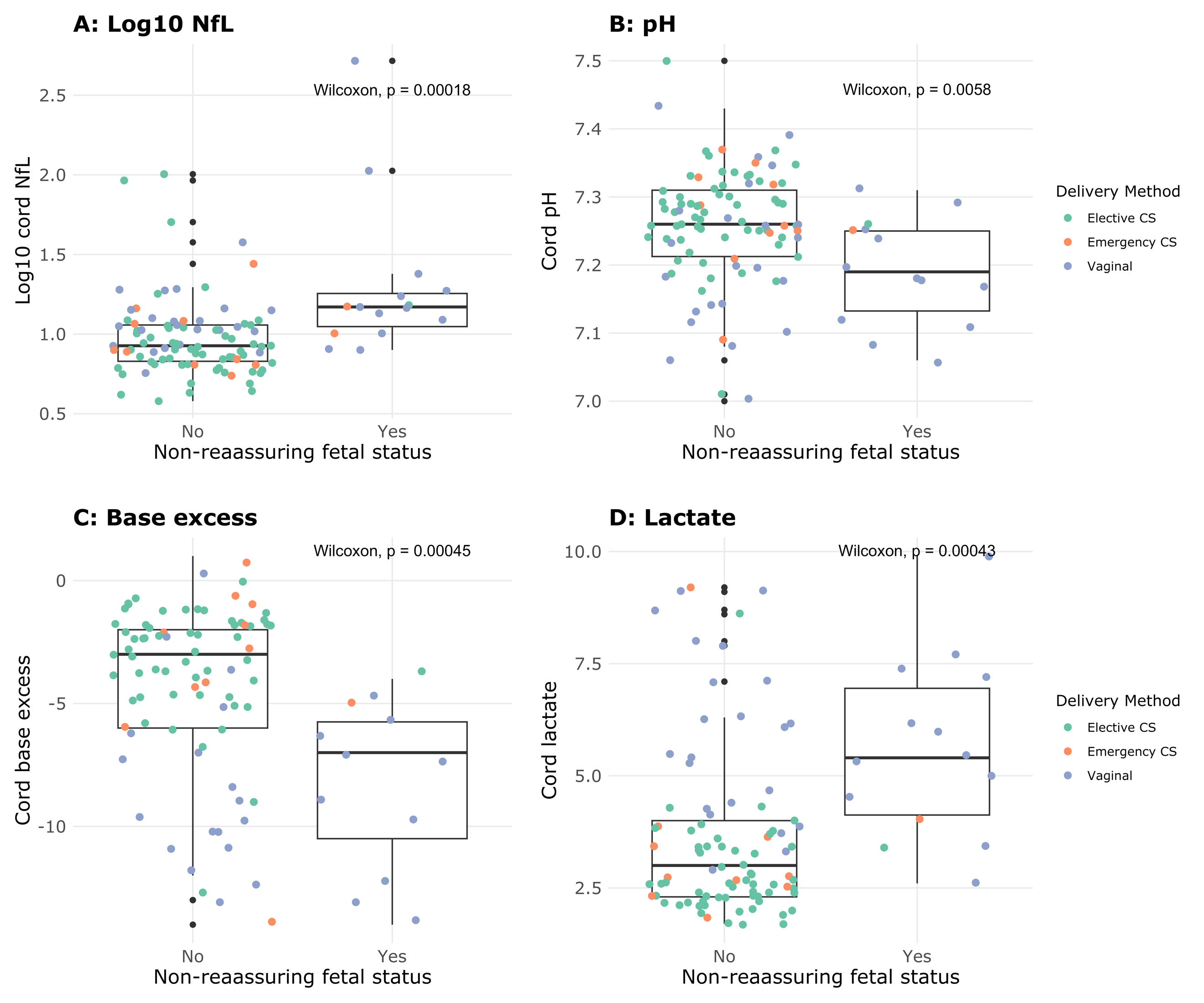

**Supplementary Figure 2.** Comparison of cord blood biomarkers levels in cases with and without documented non-reassuring fetal status. (A) NfL (n = 105), (B) pH (n = 104), (C) base excess (n = 94), and (D) lactate (n = 103).

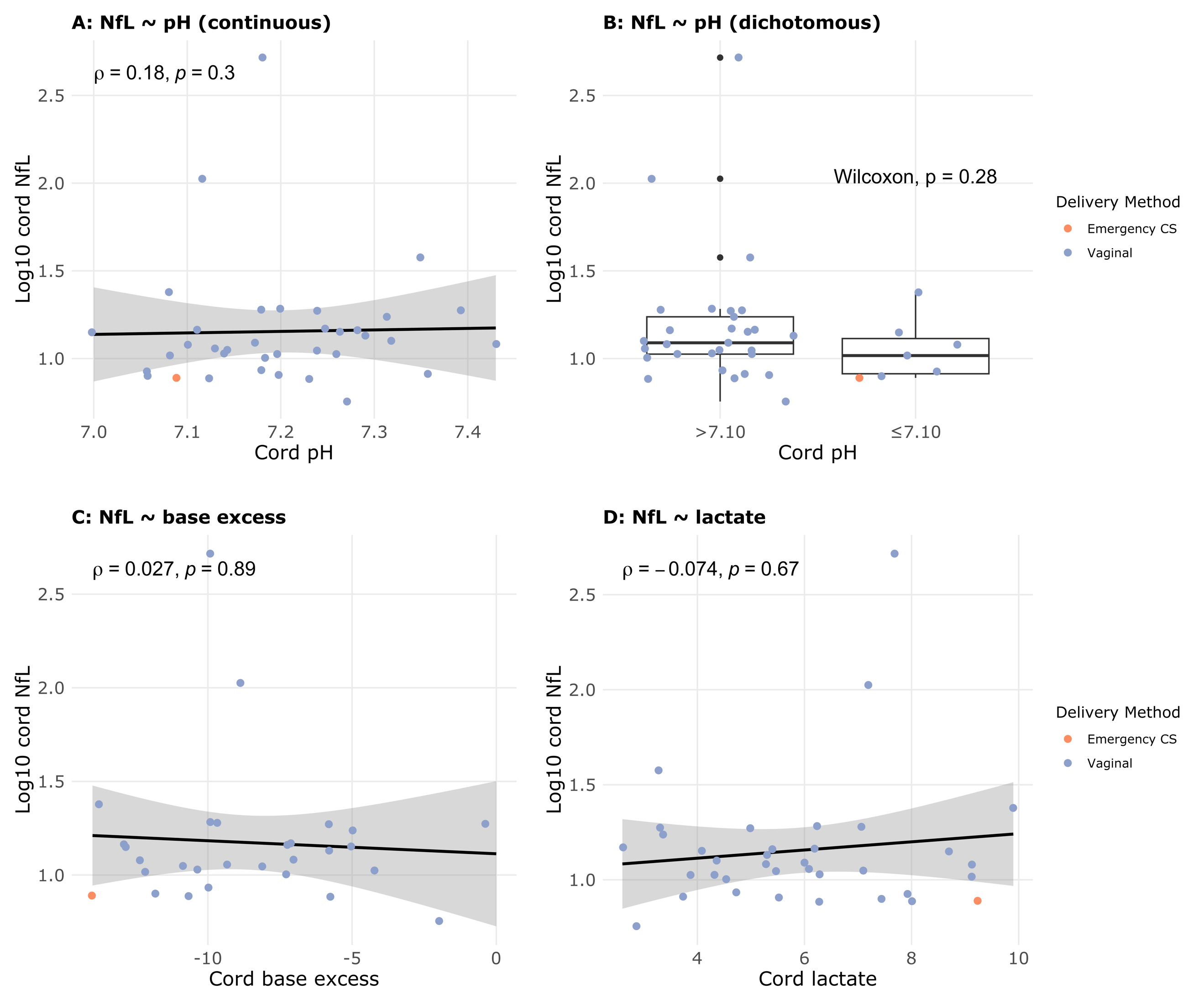

**Supplementary Figure 3.** Correlation of NfL with other biomarkers including only participants who laboured (n = 36).

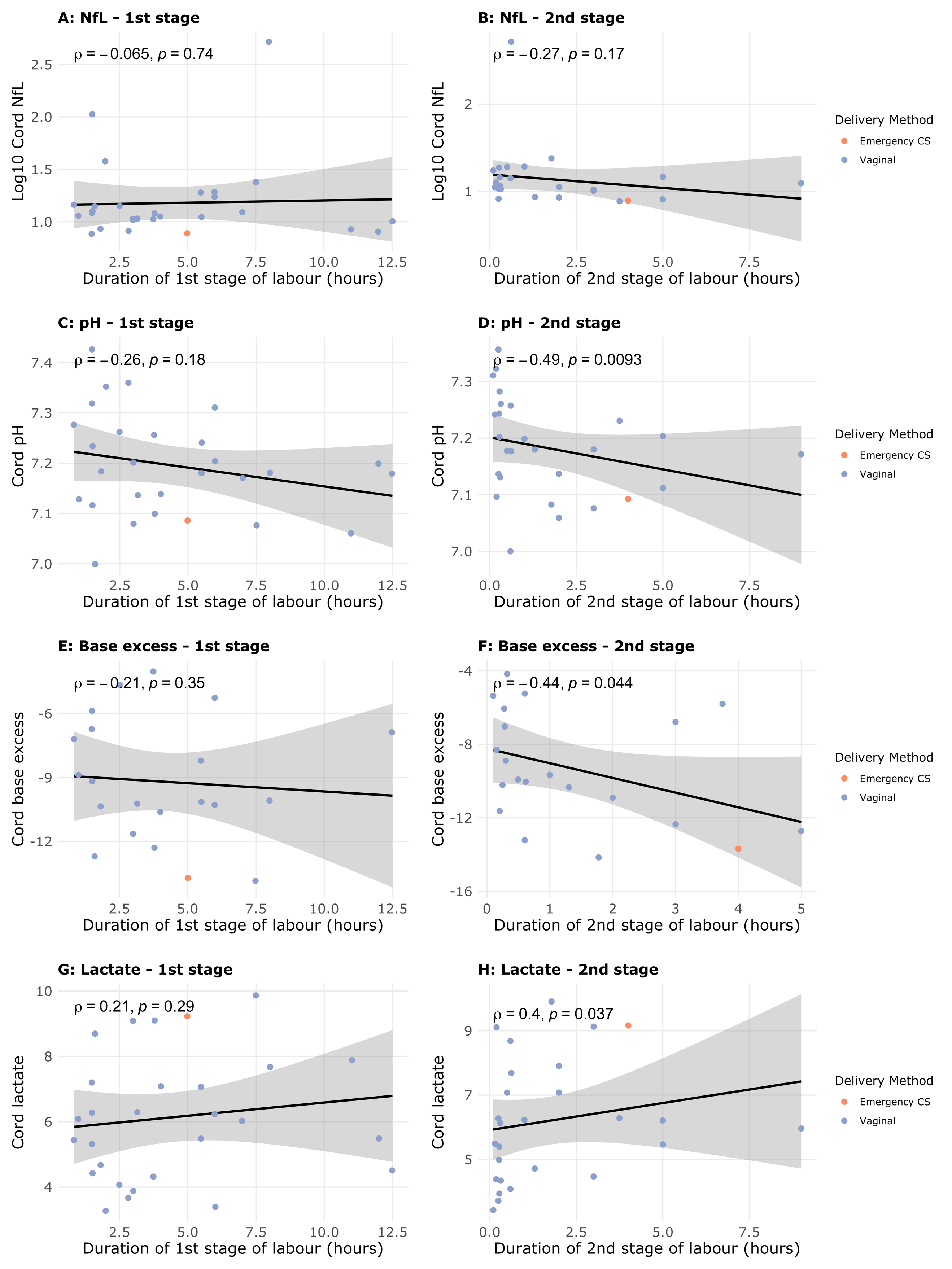

**Supplementary Figure 4.** Correlation of cord biomarkers with the duration of first and second stage of labour: (A,B) NfL, (C,D) pH, (E,F) base excess, and (G,H) lactate.

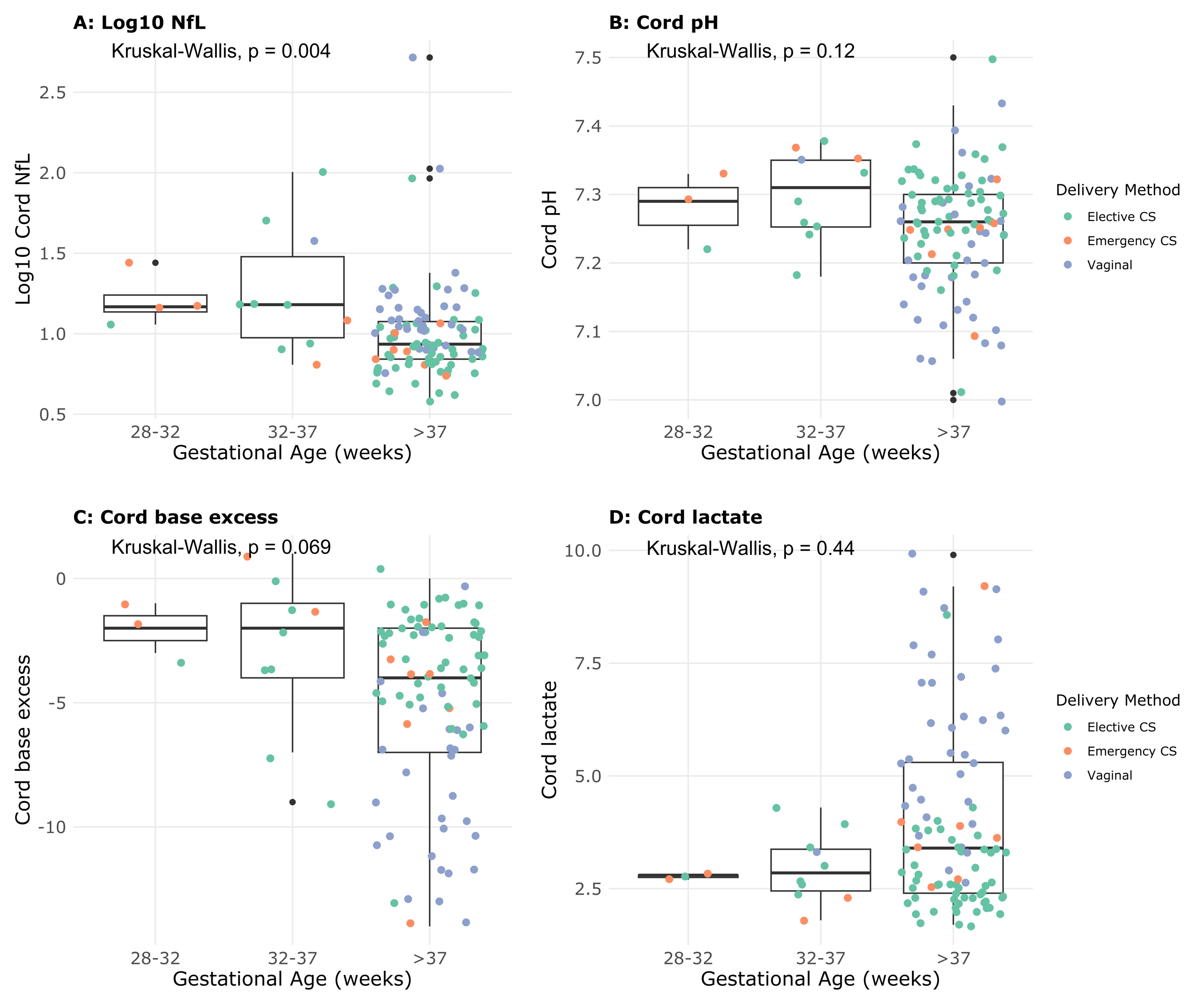

**Supplementary Figure 5.** Cord (A) NfL, (B) pH, (C) base excess, and (D) lactate concentrations by gestational age.

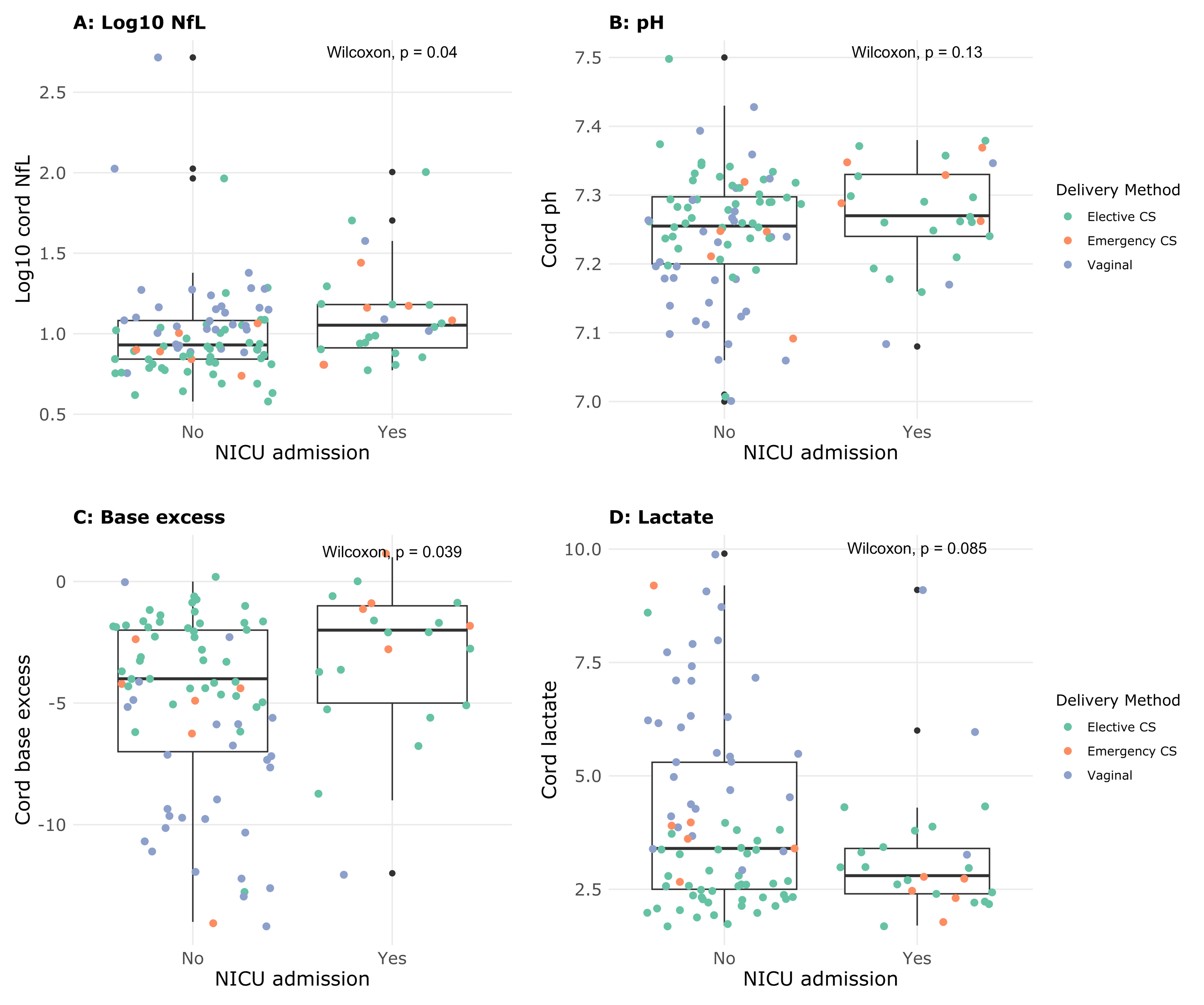

**Supplementary Figure 6.** (A) Cord NfL, (B) pH, (C) base excess, and (D) lactate in neonates who were and were not admitted to NICU.

**Supplementary Table 1.** Descriptive statistics for the study cohort, including number of missing observations.

| **Characteristic** | **N = 108***^1^* | **Missing observations** |
| --- | --- | --- |
| Antenatal Factors | | |
| Maternal age | 35 (32, 39) | 0 |
| Maternal BMI | 26 (23, 30) | 6 |
| Oligo- or polyhydramnios | 4 (3.7%) | 0 |
| Prelabour rupture of membranes | 9 (9.3%) | 11 |
| TORCH infection | 1 (0.9%) | 1 |
| IUGR | 12 (11%) | 0 |
| Maternal Factors | | |
| Diabetes | 22 (21%) | 3 |
| Anaemia | 29 (28%) | 3 |
| Smoking | 4 (3.8%) | 2 |
| Maternal infection | 10 (9.3%) | 0 |
| Pre-eclampsia | 2 (1.9%) | 0 |
| Hypertension or Pre-eclampsia | 8 (7.5%) | 1 |
| Intrapartum Factors | | |
| Gestational age (weeks) | 39.00 (37.73, 39.10) | 0 |
| Preterm birth | 14 (13%) | 0 |
| Caesarean section | 73 (68%) | 0 |
| Emergent delivery | 12 (11%) | 0 |
| Breech presentation | 8 (8.6%) | 15 |
| Instrumental delivery | 8 (7.6%) | 3 |
| Placental abruption | 1 (1.0%) | 4 |
| Oxytocin given | 25 (23%) | 0 |
| CTG abnormality | 13 (20%) | 44 |
| Non-reassuring fetal status | 16 (15%) | 3 |
| Postnatal Outcomes | | |
| APGAR ≤7 (1 min) | 14 (13%) | 0 |
| APGAR ≤7 (5 min) | 2 (1.9%) | 0 |
| NICU admission | 26 (24%) | 0 |
| Special care admission | 8 (7.4%) | 0 |
| Resuscitation at birth | 20 (19%) | 1 |
| Readmission to hospital | 4 (3.8%) | 3 |
| Anaesthetic complications | 6 (5.7%) | 2 |
| Birth-Related injuries | 10 (9.3%) | 0 |
| Neonatal sepsis | 9 (8.3%) | 0 |
| Respiratory complications | 22 (20%) | 0 |
| GIT complications | 15 (14%) | 0 |
| Haematological complications | 3 (2.8%) | 0 |
| Placental abnormalities | 11 (10%) | 1 |
| *^1^* Median (IQR); n (%) | | |

**Supplementary Table 2.** NfL association with birthweight and gestational age. NfL and lactate use an inverse Gaussian family distribution for the response variable with a log link, while pH and base excess use a Gaussian family distribution with an identity link. As base excess needed to be log-transformed, the one case with positive base excess value of 1 was changed to 0.

|  | **Log10 NfL***^1,2^* | | | **pH***^3,4^* | | | **Base excess***^5,4^* | | | **Lactate***^6,2^* | | |
| --- | --- | --- | --- | --- | --- | --- | --- | --- | --- | --- | --- | --- |
| **Characteristic** | **exp(Beta)** | **95% CI***^7^* | **p-value***^8^* | **Beta** | **95% CI***^7^* | **p-value***^8^* | **Beta** | **95% CI***^7^* | **p-value***^8^* | **exp(Beta)** | **95% CI***^7^* | **p-value***^8^* |
| **Unadjusted analysis** | | | | | | | | | | | | |
| Birthweight (kg) | 0.91 | 0.84, 0.98 | 0.017* | -0.03 | -0.06, -0.01 | 0.005** | -1.04 | -2.10, 0.02 | 0.054 | 1.10 | 0.94, 1.27 | 0.191 |
| **Adjusted analysis** | | | | | | | | | | | | |
| Birthweight (kg) | 0.89 | 0.81, 0.98 | 0.025* | -0.03 | -0.06, 0.00 | 0.084 | 0.15 | -1.17, 1.46 | 0.824 | 0.94 | 0.80, 1.11 | 0.461 |
| Gestational age (weeks) | 1.01 | 0.98, 1.04 | 0.545 | 0.00 | -0.01, 0.01 | 0.440 | -0.62 | -1.05, -0.19 | 0.005** | 1.07 | 1.02, 1.12 | 0.007** |
| *^1^* Unadjusted model: Log-likelihood = 2.12; Deviance = 6.20; AIC = 1.76; BIC = 9.81; No. Obs. = 108. Adjusted model: Log-likelihood = 2.39; Deviance = 6.17; AIC = 3.22; BIC = 13.9; No. Obs. = 108; p-value from F-test comparing unadjusted vs. adjusted model fit = 0.531 *^2^* Inverse Gaussian family models with log link. Regression coefficients correspond to the multiplicative change in the conditional mean of the biomarker concentration per 1-unit increase in the predictor *^3^* Unadjusted model: Log-likelihood = 114; Deviance = 0.744; AIC = -222; BIC = -214; No. Obs. = 107. Adjusted model: Log-likelihood = 114; Deviance = 0.740; AIC = -221; BIC = -210; No. Obs. = 107; p-value from F-test comparing unadjusted vs. adjusted model fit = 0.440 *^4^* Gaussian family models with identity link. Regression coefficients correspond to the absolute change in the conditional mean of the biomarker concentration per 1-unit increase in the predictor *^5^* Unadjusted model: Log-likelihood = -261; Deviance = 1,244; AIC = 529; BIC = 536; No. Obs. = 97. Adjusted model: Log-likelihood = -257; Deviance = 1,144; AIC = 523; BIC = 533; No. Obs. = 97; p-value from F-test comparing unadjusted vs. adjusted model fit = 0.005 *^6^* Unadjusted model: Log-likelihood = -197; Deviance = 6.04; AIC = 400; BIC = 408; No. Obs. = 106. Adjusted model: Log-likelihood = -193; Deviance = 5.57; AIC = 394; BIC = 404; No. Obs. = 106; p-value from F-test comparing unadjusted vs. adjusted model fit = 0.006 *^7^* CI = Confidence Interval *^8^* *p<0.05; **p<0.01; ***p<0.001 | | | | | | | | | | | | |

**Supplementary Table 3.** NfL association with head circumference and gestational age. NfL and lactate use a Gamma family distribution for the response variable, while pH and base excess use a Gaussian family distribution.

|  | **Log10 NfL***^1,2^* | | | **pH***^3,4^* | | | **Base excess***^5,4^* | | | **Lactate***^6,2^* | | |
| --- | --- | --- | --- | --- | --- | --- | --- | --- | --- | --- | --- | --- |
| **Characteristic** | **exp(Beta)** | **95% CI***^7^* | **p-value***^8^* | **Beta** | **95% CI***^7^* | **p-value***^8^* | **Beta** | **95% CI***^7^* | **p-value***^8^* | **exp(Beta)** | **95% CI***^7^* | **p-value***^8^* |
| **Unadjusted analysis** | | | | | | | | | | | | |
| Head circumference (cm) | 0.98 | 0.97, 1.00 | 0.084 | -0.01 | -0.01, 0.00 | 0.043* | -0.11 | -0.40, 0.18 | 0.460 | 1.01 | 0.98, 1.06 | 0.474 |
| **Adjusted analysis** | | | | | | | | | | | | |
| Head circumference (cm) | 0.99 | 0.97, 1.00 | 0.168 | 0.00 | -0.01, 0.00 | 0.294 | 0.19 | -0.13, 0.52 | 0.240 | 0.99 | 0.96, 1.02 | 0.479 |
| Gestational age (weeks) | 1.00 | 0.97, 1.02 | 0.831 | -0.01 | -0.02, 0.00 | 0.152 | -0.69 | -1.08, -0.30 | <0.001*** | 1.07 | 1.02, 1.11 | 0.006** |
| *^1^* Unadjusted model: Log-likelihood = -0.936; Deviance = 6.39; AIC = 7.87; BIC = 15.8; No. Obs. = 105. Adjusted model: Log-likelihood = -0.901; Deviance = 6.39; AIC = 9.80; BIC = 20.4; No. Obs. = 105; p-value from F-test comparing unadjusted vs. adjusted model fit = 0.822 *^2^* Inverse Gaussian family models with log link. Regression coefficients correspond to the multiplicative change in the conditional mean of the biomarker concentration per 1-unit increase in the predictor *^3^* Unadjusted model: Log-likelihood = 110; Deviance = 0.733; AIC = -214; BIC = -206; No. Obs. = 104. Adjusted model: Log-likelihood = 111; Deviance = 0.718; AIC = -214; BIC = -204; No. Obs. = 104; p-value from F-test comparing unadjusted vs. adjusted model fit = 0.152 *^4^* Gaussian family models with identity link. Regression coefficients correspond to the absolute change in the conditional mean of the biomarker concentration per 1-unit increase in the predictor *^5^* Unadjusted model: Log-likelihood = -254; Deviance = 1,220; AIC = 514; BIC = 521; No. Obs. = 94. Adjusted model: Log-likelihood = -248; Deviance = 1,075; AIC = 504; BIC = 514; No. Obs. = 94; p-value from F-test comparing unadjusted vs. adjusted model fit = 0.001 *^6^* Unadjusted model: Log-likelihood = -190; Deviance = 5.86; AIC = 387; BIC = 395; No. Obs. = 103. Adjusted model: Log-likelihood = -186; Deviance = 5.39; AIC = 380; BIC = 390; No. Obs. = 103; p-value from F-test comparing unadjusted vs. adjusted model fit = 0.005 *^7^* CI = Confidence Interval *^8^* *p<0.05; **p<0.01; ***p<0.001 | | | | | | | | | | | | |

**Supplementary Table 4**. Binomial-family regression predicting requirement for resuscitation at birth.

|  | **Log10 cord NfL***^1^* | | | **Cord pH***^2^* | | | **Cord base excess***^3^* | | | **Cord lactate***^4^* | | |
| --- | --- | --- | --- | --- | --- | --- | --- | --- | --- | --- | --- | --- |
| **Characteristic** | **OR***^5^* | **95% CI***^5^* | **p-value***^6^* | **OR***^5^* | **95% CI***^5^* | **p-value***^6^* | **OR***^5^* | **95% CI***^5^* | **p-value***^6^* | **OR***^5^* | **95% CI***^5^* | **p-value***^6^* |
| **Unadjusted model** | | | | | | | | | | | | |
| Cord biomarker (standardised) | 1.36 | 0.87, 2.12 | 0.157 | 0.88 | 0.54, 1.45 | 0.604 | 1.09 | 0.65, 1.95 | 0.766 | 0.98 | 0.56, 1.58 | 0.933 |
| **Adjusted model** | | | | | | | | | | | | |
| Cord biomarker (standardised) | 0.88 | 0.40, 1.58 | 0.720 | 0.56 | 0.30, 1.01 | 0.056 | 0.82 | 0.45, 1.57 | 0.518 | 1.31 | 0.71, 2.27 | 0.344 |
| Preterm birth |  |  |  |  |  |  |  |  |  |  |  |  |
| No | — | — |  | — | — |  | — | — |  | — | — |  |
| Yes | 47.6 | 9.51, 418 | <0.001*** | 63.9 | 12.2, 537 | <0.001*** | 38.8 | 7.83, 302 | <0.001*** | 43.8 | 9.24, 329 | <0.001*** |
| *^1^* Unadjusted model: AIC = 105; BIC = 111; No. Obs. = 107. Adjusted model: AIC = 80.4; BIC = 88.4; No. Obs. = 106 *^2^* Unadjusted model: AIC = 103; BIC = 109; No. Obs. = 106. Adjusted model: AIC = 76.5; BIC = 84.4; No. Obs. = 105 *^3^* Unadjusted model: AIC = 93.6; BIC = 98.7; No. Obs. = 96. Adjusted model: AIC = 73.3; BIC = 80.9; No. Obs. = 96 *^4^* Unadjusted model: AIC = 103; BIC = 109; No. Obs. = 105. Adjusted model: AIC = 79.1; BIC = 87.0; No. Obs. = 104 *^5^* OR = Odds Ratio, CI = Confidence Interval *^6^* *p<0.05; **p<0.01; ***p<0.001 | | | | | | | | | | | | |

**Supplementary Table 5.** Cord biomarkers as predictors of NICU length of stay. A negative binomial model is used in all cases.

|  | **Log10 NfL***^1^* | | | **pH***^2^* | | | **Base excess***^3^* | | | **Lactate***^4^* | | |
| --- | --- | --- | --- | --- | --- | --- | --- | --- | --- | --- | --- | --- |
| **Characteristic** | **IRR** | **95% CI***^5^* | **p-value***^6^* | **IRR** | **95% CI***^5^* | **p-value***^6^* | **IRR** | **95% CI***^5^* | **p-value***^6^* | **IRR** | **95% CI***^5^* | **p-value***^6^* |
| **Unadjusted analysis** | | | | | | | | | | | | |
| Cord biomarker (standardised) | 3.04 | 0.94, 9.80 | 0.063 | 3.27 | 1.65, 6.49 | <0.001*** | 3.85 | 1.79, 8.27 | <0.001*** | 0.28 | 0.12, 0.65 | 0.003** |
| **Adjusted analysis** | | | | | | | | | | | | |
| Cord biomarker (standardised) | 1.51 | 0.71, 3.19 | 0.284 | 1.10 | 0.62, 1.96 | 0.754 | 1.03 | 0.46, 2.27 | 0.951 | 1.07 | 0.46, 2.47 | 0.876 |
| Mode of delivery |  |  |  |  |  |  |  |  |  |  |  |  |
| Elective CS | — | — |  | — | — |  | — | — |  | — | — |  |
| Emergency CS | 2.12 | 0.51, 8.76 | 0.298 | 1.89 | 0.44, 8.08 | 0.389 | 1.98 | 0.43, 9.01 | 0.378 | 1.97 | 0.47, 8.26 | 0.356 |
| Vaginal | 0.34 | 0.08, 1.48 | 0.151 | 0.50 | 0.13, 1.96 | 0.319 | 0.12 | 0.01, 1.36 | 0.087 | 0.43 | 0.08, 2.33 | 0.324 |
| Gestational age | 0.60 | 0.47, 0.77 | <0.001*** | 0.57 | 0.45, 0.74 | <0.001*** | 0.52 | 0.40, 0.68 | <0.001*** | 0.56 | 0.44, 0.72 | <0.001*** |
| *^1^* Unadjusted model: AIC = 254; BIC = 262; No. Obs. = 106. Adjusted model: AIC = 229; BIC = 245; No. Obs. = 106; p-value from Chi-squared test comparing unadjusted vs. adjusted model fit = 0.000 *^2^* Unadjusted model: AIC = 249; BIC = 257; No. Obs. = 106. Adjusted model: AIC = 231; BIC = 247; No. Obs. = 106; p-value from Chi-squared test comparing unadjusted vs. adjusted model fit = 0.000 *^3^* Unadjusted model: AIC = 214; BIC = 222; No. Obs. = 96. Adjusted model: AIC = 191; BIC = 207; No. Obs. = 96; p-value from Chi-squared test comparing unadjusted vs. adjusted model fit = 0.000 *^4^* Unadjusted model: AIC = 251; BIC = 259; No. Obs. = 105. Adjusted model: AIC = 230; BIC = 246; No. Obs. = 105; p-value from Chi-squared test comparing unadjusted vs. adjusted model fit = 0.000 *^5^* CI = Confidence Interval *^6^* *p<0.05; **p<0.01; ***p<0.001 | | | | | | | | | | | | |
