## Appendices for "The association of umbilical cord blood neurofilament light with non-reassuring fetal status: a prospective observational study"

**Appendix 1:** NSW Health Intrapartum Electronic Fetal Monitoring (EFM) Staging Criteria (NSW Health, 2018).

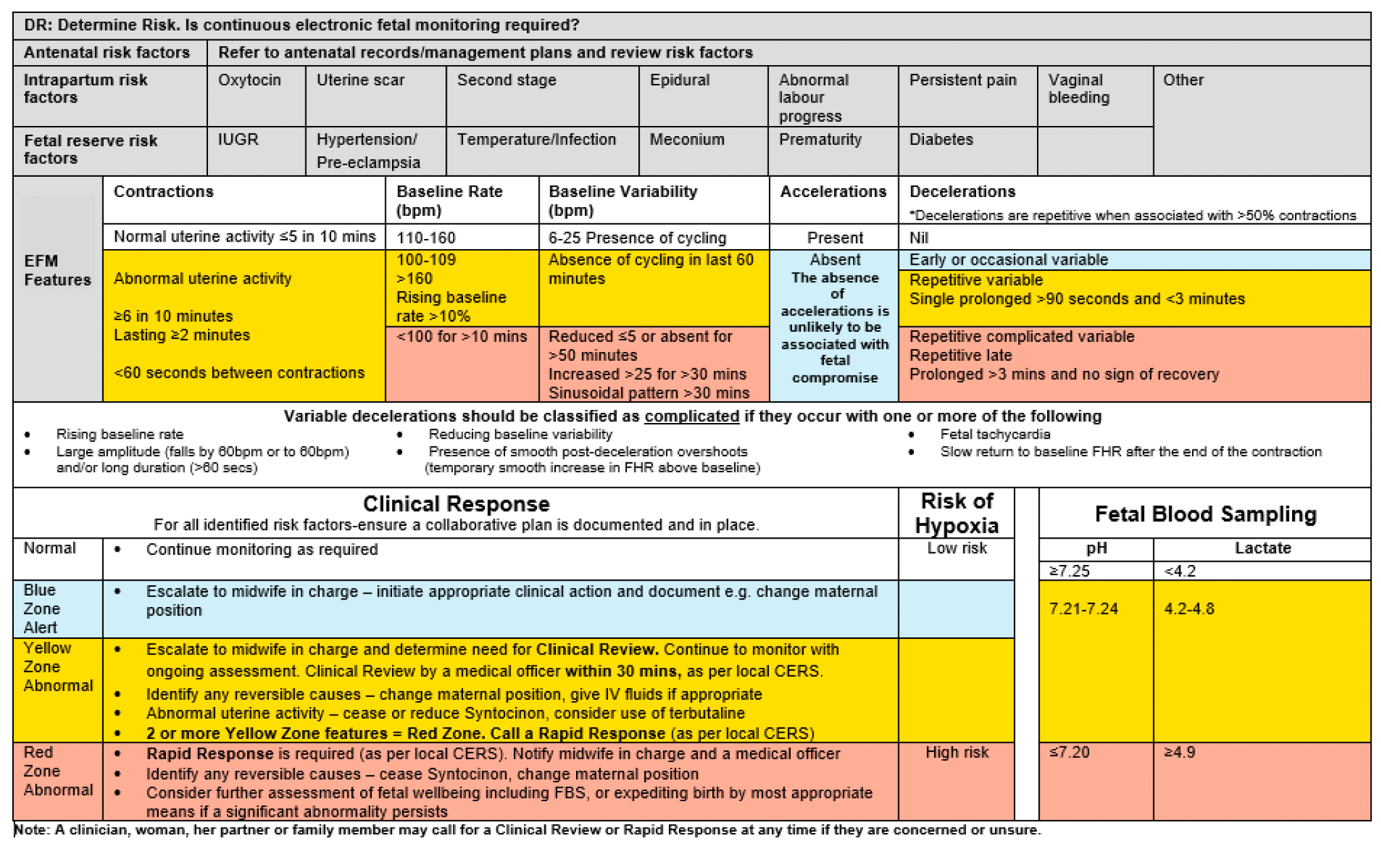

**Appendix 2.** List of variables that were extracted from the medical record.

| **Outcome Variable** | **Options** |
| --- | --- |
| Ethnicity | 1, Oceanian \| 2, North-west European \| 3, Southern/Eastern European \| 4, North African/Middle Eastern \| 5, South-East Asian \| 6, North-East Asian \| 7, Southern/Central Asian \| 8, Americas \| 9, Sub-Saharan African \| 10, Other |
| Weight |  |
| Height |  |
| Diabetes | 1, Type 1 \| 2, Type 2 \| 3, Gestational \| 4, None \| 5, Unknown |
| Multiple Pregnancies | 1, MCMA \| 2, MCDA \| 3, DCDA \| 4, No \| 5, Unknown |
| Hypertension | 1, Gestational hypertension \| 2, Pre-existing \| 3, None |
| Folate Supplement | 1, Taken regularly \| 2, Taken irregularly \| 3, Not Taken \| 4, Unknown |
| Maternal Health | 1, Cardiovascular disease \| 2, Thrombophilia \| 3, Autoimmune disease \| 4, Inflammatory Bowel Disease \| 5, Asthma \| 6, Obesity \| 7, Malignancy \| 8, Neurological disease (Description: __________________) \| 9, Other (Description: ____________________________) |
| Group B Streptococcus status | 1, positive \| 2, negative \| 3, not done \| 4, unknown |
| Pre-natal tests | 1, 1st trimester combined screening \| 2, 2nd trimester combined screening \| 3, NIPT \| 4, None \| 5, Unknown |
| Pre-natal Ultrasound | 1, Abnormal (description) \| 2, Normal \| 3, Not done \| 4, Unknown |
| Infectious status:  HIV  HBV  HCV  Syphilis  Rubella  Varicella | 1, Positive \| 2, Negative \| 3, Not Screened \| 4, Unknown |
| Midstream Urine | 1, Asymptomatic bacteriuria \| 2, UTI \| 3, Normal \| 4, Not Done \| 5, Unknown |
| Pre-eclampsia | 1, Pre-eclampsia \| 2, Eclampsia \| 3, HELPP \| 4, None \| 5, Unknown |
| Placental Abnormalities | 1, Subchorionic haematoma \| 2, placenta previa \| 3, vasa praevia \| 4, single umbilical artery \| 5, velamentous cord insertion \| 6, circumvallate \| 7, other \| 8, none \| 9, unknown |
| Maternal embolism | 1, thromboembolism \| 2, air embolism \| 3, amniotic fluid embolism \| 4, none \| 5, unknown |
| Intrauterine growth restriction | 1, Small for gestational age \| 2, Fetal Growth Restriction \| 3, None \| 4, Unknown |
| Prelabour rupture of membranes | 1, PROM \| 2, PROM > 24 hours from birth \| 3, None \| 4, N/A \| 5, Unknown |
| Preterm birth reason | 1, Spontaneous \| 2, Iatrogenic \| 3, Other \| 4, Unknown |
| Induction Type | 1, AROM \| 2, Cervical Ripening \| 3, Hormones \| 4, Other |
| Duration of stage 1  Duration of stage 2 |  |
| Fetal presentation | 1, Cephalic \| 2, Breech \| 3, Transverse \| 4, Oblique \| 5, Unstable \| 6, Fundic/Cord Presentation \| 7, Unknown |
| Pain management | 1, Epidural/Spinal \| 2, Nitrous oxide \| 3, Pethidine \| 4, Other \| 5, None \| 6, Unknown |
| Caesarean section delivery | 1, Cat 1 - Details: \| 2, Cat 2 \| 3, Cat 3 \| 4, Cat 4 \| 5, Elective \| 6, N/A \| 7, Unknown |
| Type of Instrumental Delivery | 1, Vacuum (kiwi cup) \| 2, Vacuum (mityvac) \| 3, Vacuum (not specified) \| 4, Forceps \| 5, Other |
| Uterotonic agent administration | 1, Oxytocin \| 2, Ergometrine \| 3, Syntometrine \| 4, Misoprostol \| 5, Carboprost \| 6, Carbetocin |
| Complications during birth | 1, Uterine rupture \| 2, Umbilical cord prolapse \| 3, Chorioamnionitis \| 4, Puerperal Sepsis \| 5, Other \| 6, None \| 7, Unknown |
| Birth Weight |  |
| Gestational Age |  |
| Head circumference |  |
| Injuries from birth | 1, Cephalohaemoatoma \| 2, Subgaleal haemorrhage \| 3, Caput succedaneum \| 4, Erbs Palsy \| 5, Clavicle/humeral fracture \| 6, Cranial nerve palsy \| 7, Laceration \| 8, Facial Bruising \| 9, Other (describe) \| 10, None \| 11, Unknown |
| Cord Blood lactate  Cord Blood base excess |  |
| Cord blood - pH |  |
| NICU/Special Care Duration of Stay |  |
| Apgar at 1 and 5 minutes |  |
| Torch infections | 1, Toxoplasma \| 2, Rubella \| 3, Cytomegalovirus \| 4, HIV \| 5, Hepatitis Viruses \| 6, Herpes simplex \| 7, Syphilis \| 8, Varicella zoster virus \| 9, Parvovirus B19 \| 10, None \| 11, Unknown |
| Genetic Disorders | 12, Trisomy 21 \| 13, Trisomy 18, 14, 13 \| 14, Klinefelter Syndrome \| 15, Turner syndrome \| 16, Spinal Muscular Atrophy \| 17, Fragile x syndrome \| 18, Cystic Fibrosis \| 19, Other \| 20, None \| 21, Unknown |
| Birth Defects | 1, Congenital heart disease \| 2, neural tube defects- description \| 3, Abdominal wall defects \| 4, Congenital diaphragmatic hernia \| 5, Tracheal abnormalities \| 6, Oesophageal atresia \| 7, Pulmonary Hypoplasia \| 8, Other - describe \| 9, None \| 10, Unknown |
| Neonatal Sepsis | 1, Early onset \| 2, Late onset \| 3, None \| 4, Unknown. |
| Metabolic Complications | 1, Hypoglycaemia \| 2, Hypothermia \| 3, Anaemia \| 4, Polycythaemia \| 5, Congenital Hypothyroidism \| 6, Metabolic bone disease of prematurity \| 7, Haemolytic disease of the newborn \| 8, Congenital adrenal hyperplasia \| 9, OTHER - description \| 10, None \| 11, Unknown |
| Gastrointestinal Complications | 1, Necrotising enterocolitis \| 2, Jaundice \| 3, Biliary atresia \| 4, Neonatal hepatitis \| 5, Other (Description:_________________________) \| 6, None \| 7, Unknown |
| Respiratory Complications | 1, Respiratory distress syndrome \| 2, Apnoea of prematurity \| 3, Bronchopulmonary dysplasia \| 4, Transient tachypnoea of the newborn \| 5, Pneumothorax \| 6, Pneumonia \| 7, Meconium aspiration syndrome \| 8, Persistent pulmonary hypertension of the newborn \| 9, Other (Description:____________________________) \| 10, None \| 11, Unknown |
| Neurological Complications | 1, Cerebral palsy \| 2, Intraventricular haemorrhage \| 3, Retinopathy of prematurity \| 4, Hypoxic-ischemia encephalopathy \| 5, Periventricular leukomalacia \| 6, Seizures \| 7, Movement disorders \| 8, Other (Description:_________________________) \| 9, None \| 10, Unknown |
| Haematological Complications | 1, Anaemia of prematurity \| 2, Physiological anaemia of the newborn \| 3, Other \| 4, None \| 5, Unknown |
| Injuries from Birth | 1, Cephalohematoma \| 2, Subgalea haemorrhage \| 3, Caput succedaneum \| 4, Erbs Palsy \| 5, Clavicle/humeral fracture \| 6, Cranial nerve palsy \| 7, Laceration \| 8, Facial Bruising \| 9, Other (describe) \| 10, None \| 11, Unknown |
